# Associations of exercise and social support with mental health during quarantine and social-distancing measures during the COVID-19 pandemic: A cross-sectional survey in Germany

**DOI:** 10.1101/2020.07.01.20144105

**Authors:** Leonie Louisa Bauer, Britta Seiffer, Clara Deinhart, Beatrice Atrott, Gorden Sudeck, Martin Hautzinger, Inka Rösel, Sebastian Wolf

## Abstract

**Introduction:** Social distancing and quarantine measures applied during the COVID-19 pandemic might result in mental health problems. In this cross-sectional study we examined if perceived social support, exercise in minutes per week and change in exercise are protective factors regarding symptoms of depression, anxiety, and sleeping disorders.

**Method:** In April 2020, n = 4271 German adults completed an online survey including mental health questionnaires regarding depression (PHQ-D), anxiety (PHQ-D) and sleep (PSQI), as well as questionnaires related to protective factors such as exercise (BSA-F), physical activity-related health competence (PAHCO) and social support (F-SozU).

**Results:** Complete case analysis (n = 3700; mean age 33.13 ± 11.73 years, 78.6 % females) resulted in elevated prevalence of depressive disorder (31.4%), panic disorder (5.7%) and other anxiety disorders (7.4%). 58.3% reported symptoms of insomnia. Three separate models of multiple regression were conducted. Perceived social support was associated with lower values of anxiety (*beta* = −0.10; *t*(19) = −6.46; *p* >0.001), lower values of depressive symptoms (*beta* = −0.22; *t*(19) = −15.71; *p* < .001) and lower values of sleeping disorder symptoms (*beta* = −0.15; *t*(19) = −9.55; *p* < .001). Change towards less exercise compared to the time before Covid-19 was associated with and higher values of anxiety (*beta* = −0.05; *t*(19) = −2.85; *p*= .004), higher values of depressive symptoms (*beta* = −0.08; *t*(19) = - 5.69; *p <* .001), and higher values of sleeping disorder symptoms (*beta* = −0.07; *t*(19) = −4.54; *p* < .000). Post-hoc analysis (ANOVAs) revealed that a change towards less exercise was significantly associated with more depressive, anxiety and sleeping disorder symptoms whereas a positive change was not. No significant association was found for exercise in minutes per week for all outcomes.

**Conclusion:** The COVID-19 pandemic seems to have a negative impact on mental health in the German population. Social Support and a stable amount of exercise might attenuate these negative mental health consequences. Ongoing monitoring of the impact of the pandemic on mental health and possible protective factors is needed in order to create a basis for the development of appropriate prevention and intervention measures.

## 1. Background

With 8327805 confirmed cases all over the world (up to June 19th, 2020; World Health Organization, 2020), the COVID-19 disease is a global public health emergency. The World Health Organization (WHO) has issued recommendations to implement social distancing measures for the general public as well as quarantining procedures, for people infected with the COVID-19 disease.

Similar measures have been enforced during recent pandemic, such as the 2003 outbreak of SARS and the 2014 outbreak of Ebola. As a result, studies reported increased symptoms of post-traumatic-stress disorders and depressive disorders, as well as a 30% increase in suicide rates in populations concerned by the measures. In addition, they reported increased prevalence of sleep disturbances, confusion, and anger (Brooks et al., 2020; Lee et al., 2007; Yip, Cheung, Chau, & Law, 2010).

In line with this evidence, studies investigating the impact of social distancing and quarantine measures on mental health consequences in countries severely affected by Covid-19 (China, United States of America, Brazil, and Italy) report high overall prevalence of symptoms of mental illness. Prevalence of depressive symptoms ranged from 17.3% to 43.6%. 7.1% to 45.4% of participants reported raised anxiety symptoms. 7.3% to 18.2% of participants reported reduced sleep quality and 31.1% of participants reported raised levels of perceived stress (Cao et al., 2020; Huang & Zhao, 2020; Liu, Zhang, Wong, & Hyun, 2020; Meyer et al., 2020; Rossi et al., 2020; Schuch et al., 2020; C. Wang et al., 2020).

In May 2020, 2.02 million employees in Germany were faced with short-time working arrangements due to economic consequences of the pandemic. The unemployment rate has risen 25% compared to the previous year, leading to financial insecurity (Federal Employment Agency, 2020). With 188534 confirmed infections and 8872 deaths due to COVID-19 in Germany, many people have been affected by serious illness or the loss of friends and family members (up to June 19th, 2020; World Health Organization, 2020). These negative life events are correlated to higher depression and anxiety symptoms (Dalgard et al., 2006; Miloyan, Joseph Bienvenu, Brilot, & Eaton, 2018). In addition, unemployment, and the recommendation of working from home might have led to a loss of daily structure and positive reinforcement, which in turn might be associated with poorer mental health (Paul, Geithner, & Moser, 2009). Further the implementation of social distancing and quarantine procedures might have led to loneliness and perceived lack of social support, which is associated with raised depression and anxiety symptoms (Santini et al., 2020).

Evidence from meta-analyses and large cohort studies indicates the importance of social support as a protective factor for depression (Gariepy, Honkaniemi, & Quesnel-Vallee, 2016; Grav, Hellzen, Romild, & Stordal, 2012; X. Wang, Cai, Qian, & Peng, 2014). Regarding exposure to negative life events, Dalgard et al. (2006) report symptoms of depression to increase significantly with decreasing social support. In the context of the COVID-19 pandemic, social support was found to be negatively associated with anxiety (Cao et al., 2020) and depression (Liu et al., 2020) and to correlate positively with sleep quality (Xiao, Zhang, Kong, Li, & Yang, 2020).

According to recent meta-analyses, physical activity (PA) could be a protective factor for mental illnesses (Harvey et al., 2018; Schuch et al., 2019; Schuch et al., 2018). Indeed, Schuch et al. (2020) reported that participants, who engaged in 30 minutes or more of moderate to vigorous PA (MVPA) had decreased odds of depressive or anxiety symptoms during the COVID-19 pandemic. In addition, Meyer et al. (2020) reported that compared to participants that kept adhering to MVPA (according to physical activity guidelines; American College of Sports Medicine, 2017) during COVID-19 measures, those who decreased their levels of MVPA reported more depressive symptoms, loneliness, and stress as well as lower positive mental health.

In their model of PA-related Health Competence (PAHCO), Carl, Sudeck, and Pfeifer (2020) describe the need of major competences in order to adhere to daily PA. One especially important in the context of mental health might be PA-specific affect regulation. Sudeck and Pfeifer (2016) assumed that not just the quantity and intensity of PA, but also specific self-regulation skills are important in order to reach optimal effects on mental health. Indeed, Sudeck, Jeckel, and Schubert (2018) showed that people who engage in PA with the scope and the skill to regulate their (negative) affective states show greater benefits in affective well-being (Sudeck & Pfeifer, 2016).

In this cross-sectional study we assessed, if exercising and perceived social support attenuate the negative mental health consequences induced by social distancing and quarantine measures within the German population.

The following hypotheses are tested:

1. The changes induced by social distancing and quarantine measures are associated with higher symptoms of depression, anxiety, panic, and sleeping disorders.
2. Exercise is negatively associated with symptoms of depression, anxiety, and sleeping disorders.
3. Change towards more exercise is negatively associated with symptoms of depression, anxiety, and sleeping disorders.
4. Perceived social support is negatively associated with symptoms of depression, anxiety, and sleeping disorders.
5. Higher PA related affect regulation competence enhances the effects of physical exercise on symptoms of depression, anxiety, and sleeping disorders.

## 2. Method

In April 2020, a longitudinal online survey with three measurement points and within-subject design was commenced. Data for the first measurement point (T1) was collected during complete lockdown (April 8^th^, 2020 - April 26^th,^ 2020). Since March 16^th^, 2020, schools, non-essential businesses, sports and entertainment venues were closed. As of March 23^rd^, 2020, public gatherings of more than two persons not living in the same household were banned (RKI, 2020). The second measurement point (T2) was scheduled as these restrictions were eased (Mai, 22^nd^, 2020 – Mai, 29^th^, 2020). The last measurement point (T3) will be scheduled once the restrictions are stopped (vaccine availability). This report focuses on the data of the first measurement point (T1), resulting in a cross-sectional design. The study was approved by the local ethics committee for research at the Faculty of Economics and Social Sciences and registered on the German Clinical Trial Register (DRKS00021791).

### 2.2 Procedure

Participants were directed to the online survey software SoSci Survey (Leiner, 2016). Before starting the survey, participants provided informed consent. Participants then self-reported demographic information, COVID-19-related changes in their daily life, mental health questionnaires regarding depression, anxiety, and sleep, as well as questionnaires related to protective factors such as exercise, PA-related health competence, social support and day structure. After completing the questionnaire, participants were provided with contact numbers of phone-based counselling concerning psychological burden and domestic abuse. Participants who were interested in participating at the second and third measurement point indicated their e-Mail address at the end of the survey. All questionnaires used a self-report format.

### 2.3 Measures

#### 2.3.1 Diagnosis of Disorders

##### PHQ-D

To measure depression, panic disorders, and anxiety disorders the Patient Health Questionnaire (PHQ-D; Löwe, Spitzer, Zipfel, & Herzog, 2002) was used. Depressive symptoms (major depression and other depressive disorders) are measured with 9 items on a four-point Likert-type scale over the last two weeks. Higher scores indicate a higher intensity. The sum of all items represents the total score (range: 0-27). Individuals are classified to the degree of severity: absence of depressive disorder (0-4), mild degree of severity (5-10), medium major depression (10-14), severe major depression (15-19) and most severe major depression (20-27). The PHQ-D additionally assesses panic disorders and other anxiety disorders. Within 15 items participants are asked to indicate whether they experienced a panic attack within the last four weeks and whether certain symptoms (e.g. sweating, feeling like suffocating, fear of dying) were present during their worst panic attack. Other anxiety disorders (genialized anxiety disorders, not specified anxiety disorders) are measured with seven items, rating the severity of anxiety symptoms within the last four weeks on a three-point Likert-type scale. Higher scores indicate higher severity. Panic disorders and other anxiety disorders are evaluated dichotomously and the presence for each disorder was determined for each participant. The scale measuring depressive symptoms has an internal consistency of Cronbach’s α=.88 (Gräfe, Zipfel, Herzog, & Löwe, 2004). In this study the scale for depressive symptoms revealed an internal consistency of Cronbach’s α=.86.

##### PSQI

With 19 Items the Pittsburgh Sleep Quality Index (PSQI; Wittchen et al., 2001) assesses the sleep quality within the last four weeks using a global score consisting of the components sleep duration, sleep disturbance, sleep latency, daytime dysfunction, sleep efficiency, overall sleep quality and sleep medication use. Each component ranges from zero to three. The global score consists of all seven components and ranges from 0 to 21. Higher scores indicate lower sleep quality. Individuals are classified as not fulfilling symptoms of sleeping disorders (score 0-5) and fulfilling symptoms of sleeping disorders (6-21) (Buysse, Reynolds, Monk, Berman, & Kupfer, 1989). Previous studies reported a reliability of Cronbach’s α = .77 for the total score (Doi, Minowa, Uchiyama, & Okawa, 2001). In this study the total score revealed an internal consistency of Cronbach’s α=.69.

#### 2.3.2 Criterion Variables: Depressive, anxiety and sleeping disorder symptoms

##### PHQ-D

Depressive symptoms (major depression and other depressive disorders) were measured with 9 items on a four-point Likert-type scale, using the PHQ-9 module (module 2) of the PHQ-D (PHQ-D; Löwe, Spitzer, Zipfel, & Herzog, 2002). The sum of all items represents the total score (range: 0-27). Other anxiety symptoms were assessed using module 5, item a of the PHQ-D, rating the severity of anxiety symptoms within the last four weeks on a three-point Likert-type scale. Higher scores indicate higher severity (range: 0-2).

##### PSQI

Sleeping disorder symptoms were assessed through the global score of the PSQI (range: 0-21; see paragraph 2.3.1; Buysse et al., 1989)

#### 2.3.3 Predictor variables: Exercise, Change in Exercise, Social Support, PA-related affect regulation

##### BSA-F

The Physical Activity, Exercise, and Sport Questionnaire (BSA-F; Fuchs, Klaperski, Gerber, & Seelig, 2015) examines PA in employment, leisure time and exercise activity over the past four weeks with 17 items. In validation studies, the exercise activity index was significantly correlated to performance measures, such as maximum oxygen intake, while the PA index was only weakly correlated to those performance measures (Fuchs, Klaperski, Gerber, & Seelig, 2015). We therefore decided to include the exercise index in the analyses. To measure exercise activity, the participants could indicate the frequency (on how many days during the last four weeks) and duration (for how many minutes) of three different kind of exercise. To calculate the total score for exercise the frequency and duration of each kind of exercise was multiplied and the sum was formed. This sum was then divided by four in order to reflect the amount of physical exercise per week (in minutes/week) (Fuchs et al., 2015). Higher scores reflect more exercise activity.

##### Change in Exercise

Change in exercise was assessed by one item. Participants indicated their changes in their exercise activity, ranging from less active to more active on a seven-point Likert-type scale. For descriptive and Post-hoc analysis participants were categorized into being less active (score 1-3), just as active (score 4) and more active than before the Covid-19 pandemic (5-7).

##### F-SozU

The Perceived Social Support Questionnaire (F-SozU; Fydrich, Sommer, Tydecks, & Brähler, 2009) assesses subjectively perceived and anticipated support from the social environment. The short version consists out of 14 items rated on a five-point Likert-type scale. The mean of all items represents the total score, higher scores indicate more perceived and anticipated social support (range: 1-5). The questionnaire shows an internal consistency of Cronbach’s α = 0.94 (Fydrich, Sommer, Tydecks, & Brähler, 2009). In this study the questionnaire revealed an internal consistency of Cronbach’s α= .93.

##### PAHCO

The PA-related affect regulation subscale of the PA-related health competence questionnaire (PAHCO; Sudeck & Pfeifer, 2016) examines PA-related affect regulation with 4 items on a five-point Likert-type Scale, higher scores indicating higher competencies. The reliability of this scale was determined by Sudeck and Pfeifer (2016) with Cronbach’s α ranging from .88-.89. In this study the subscale PA-related affect regulation revealed an internal consistency of Cronbach’s α= .91.

#### 2.3.4 Covariates: Sociodemographic Information, COVID-Specific Information

##### Sociodemographic questionnaire

Participants self-reported age, gender, relationship status (single, in a steady relationship), education, whether they had children, how many of their children were under 18, and whether at least one child lived in the same household as them.

##### Covid-19 questionnaire

Participants self-reported how the Covid-19 pandemic changed their daily life. More precise, they reported information on their health status (i.e. being infected with Covid-19, being chronically or acutely ill, being unable to exercise due to chronic illness) in a dichotomous answer format (yes/no). They reported their daily structure before and during the COVID-19 pandemic (visual analogue scale (VAS); unstructured to structured), the amount of social interactions before and during the COVID-19 pandemic (VAS; few social contacts to many social contacts), their satisfaction with their social life (VAS; not satisfied to very satisfied) as well as the amount of people in their household (open answer format). We assessed information on the current job situation with items concerning change in employment situation (VAS; did not change to changed a lot), working from home (VAS; never to always), and being threatened by negative life events (multiple choice: short-time work, loss of job, insolvency, not threatened by negative life event). In addition, worries about the financial situation and supply situation (VAS; no worries to a lot of worries) were assessed in two items.

### 2.4 Data preparation and analysis

The descriptive statistics, including frequencies and percentages, were generated for categorical variables; means and standard deviations (SD) were generated for continuous variables.

Due to the small number of total missing data, we performed complete case analyses for this preprint. For the final publication imputed data with multiple imputation procedures will be presented in addition to the complete case analysis.

Three separate models of multiple regression were conducted, all including exercise, change in exercise (continuous variable), social support, and PA-related affect regulation as predictor variables, as well as the interaction effect of exercise with PA-related affect regulation. Due to skewed data distribution, the exercise variable was z-standardized. To perform a moderation analysis and compute simple slope analysis, the PA-related affect regulation variable was centered. The criterion variables for the three separate models were depressive symptoms (analysis 1), anxiety symptoms (analysis 2), and sleeping disorder symptoms (analysis 3), respectively. In the case of a significant association between change of exercise and a criterion variable, post-hoc ANOVAs with the categorial variable (less active vs. as active as before vs. more active) as independent variable was performed to determine, which change exactly influenced depressive, anxiety and sleeping disorder symptoms.

Covariates included age, gender, relationship status, suffering from a chronic disease, change in employment situation, financial worries, worries about the supply situation, satisfaction with social life, daily structure. Data preparation and statistical analyses were carried out using the Statistical Package for Social Sciences (SPSS, version 26). All p-values were two-sided, the statistical significance level was set at p <0.05.

## 3. Results

### 3.1 Missing analyses

The percentage of missing values ranged from 0 percent for some demographic and clinical variables to as high as four percent for data about changes due to the corona pandemic. 86.63% of 4271 eligible subjects completed the questionnaire without missing data. The percentage of missing data in total was 0.21%. Data were primarily missing due to item nonresponse. A Missing Values Analysis indicated that Little’s (1988) test of Missing Completely at Random (MCAR) was significant, χ2(15047, *n* = 4271) = 17488.86, *p* < .001.

### 3.2 Participants/ Inclusion criteria

4343 participants completed the questionnaire. 72 participants were excluded because they indicated to be younger than 18 years or older than 99 years old. As analyses were conducted for complete cases only, 571 participants (13.37 %) were excluded because of missing data. This left a total of n = 3700 participants. Figure 1 illustrates the reasons participants were excluded.

**Figure 1:**
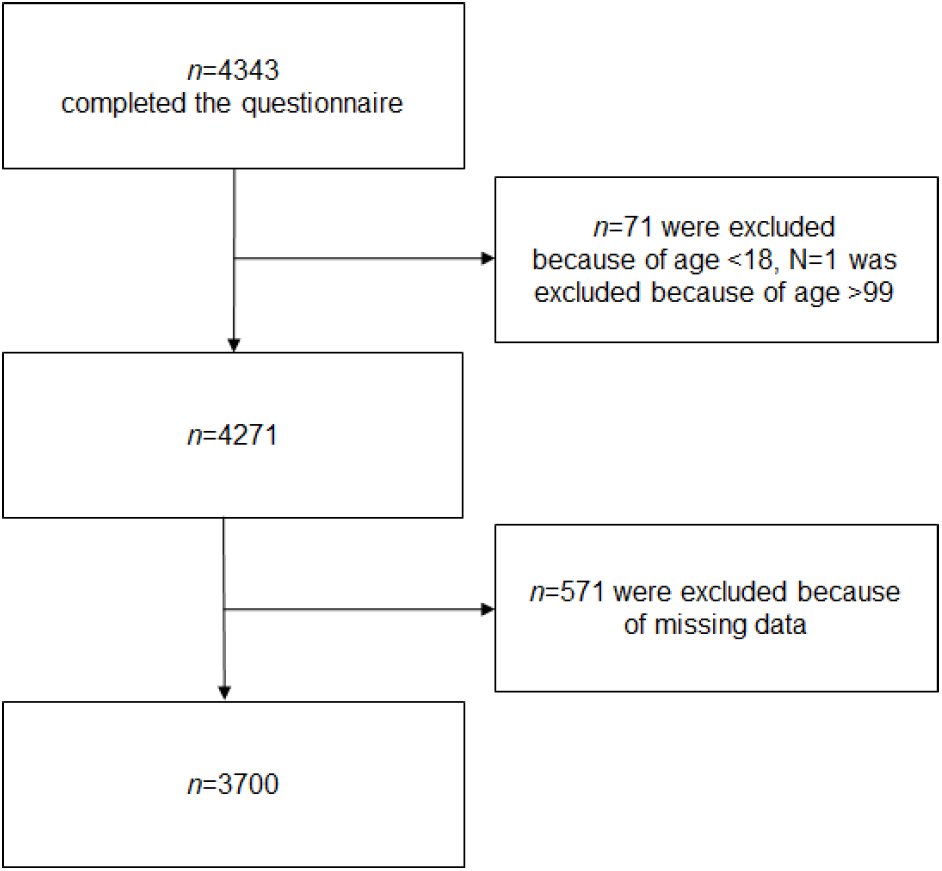
Exclusion criteria for participants

The age varied between 18 and 85 years (*M* = 33.13, *SD* = 11.73), participants were predominantly female (n = 2909; 78.6%). Participants spent most of their day time at home (*M* = 19.601 hours, *SD* = 4.432). Table 1 provides an overview of sample characteristics.

**Table 1:** Sample Characteristics

|  | *n* | [%] |
| --- | --- | --- |
| **Gender** |  |  |
| Male | 776 | 21.0 |
| Female | 2909 | 78.6 |
| Diverse | 15 | 0.4 |
| **Marital Status** |  |  |
| Single | 1240 | 33.5 |
| Relationship | 1332 | 36.0 |
| Married | 987 | 26.7 |
| Divorced | 119 | 3.2 |
| Widowed | 22 | 0.6 |
| **Amount of kids** |  |  |
| 0 | 3029 | 81.9 |
| 1 | 326 | 8.8 |
| 2 | 276 | 7.5 |
| ≥ 3 | 69 | 1.9 |
| **Academic Status** |  |  |
| No Graduation | 6 | 0.2 |
| Lower Secondary School Certificate | 37 | 1.0 |
| General Certificate of Secondary Education | 188 | 5.1 |
| High School Graduation | 933 | 25.2 |
| Professional Training | 454 | 12.3 |
| Bachelor | 755 | 20.4 |
| Master/ Diploma | 1155 | 31.2 |
| PhD | 172 | 4.6 |
| **Covid-19 diagnosis** |  |  |
| Yes | 14 | 0.4 |
| No | 3686 | 99.6 |
| **Chronic disease** |  |  |
| Yes | 329 | 8.9 |
| No | 3371 | 91.1 |
| **Threatened by short-time work, job loss, insolvency** |  |  |
| Yes | 942 | 25.5 |
| No | 2758 | 74.5 |
| **Change in Exercise** |  |  |
| Negative change | 1258 | 34.0 |
| No change | 1022 | 27.6 |
| Positive change | 1420 | 38.4 |
|  | *n* | *Mean (SD)* |
| **Age** | 3700 | 33.13 (11.73) |
| **Exercise (minutes/ week)** | 3700 | 181.33 (233.59) |
| **Sleep duration (hours/ night)** | 3700 | 8.64 (1.21) |

### 3.3 Prevalence of mental disorders

The prevalence of depressive disorders (PHQ-9 score ≥ 10, indicating a medium, severe or most severe depressive disorder) in our sample was 31.4%. This indicates a prevalence that is nearly four times as high as the prevalence of depressive disorders diagnosed using the PHQ-9 in German adults in 2013, which was 8.1% (Busch, Maske, Ryl, Schlack, & Hapke, 2013). In our sample, only 41.7% of participants indicated a good sleep quality, whereas 58.3% reported symptoms of insomnia (PSQI total score > 5). 5.7% fulfilled diagnostic criteria for panic disorder, which is also elevated compared to the prevalence in the general population in 2014, which was 2% (Jacobi et al., 2014). Table 2 provides an overview of sample scores in depressive disorder, panic disorder, other anxiety disorder and insomnia compared to prevalence in Germany.

**Table 2:** Prevalence of mental disorders in the sample of the present study during COVID-19 compared to prevalence in the general population before COVID-19

|  | Prevalence in the sample of the present study during COVID-19 |  |  |  |  |  |  |  | Prevalence in the general population before COVID-19 |  |  |
| --- | --- | --- | --- | --- | --- | --- | --- | --- | --- | --- | --- |
|  | Male (m) |  | Female (f) |  | Diverse (d) |  | Total (t) |  | m | f | t |
|  | n | % | n | % | n | % | n | % | % | % | % |
| **Depressive disorder (PHQ-D)** |  |  |  |  |  |  |  |  |  |  |  |
| *Absence (0-4)* | 324 | 41.8 | 744 | 25.6 | 3 | 20.0 | 1071 | 28.9 |  |  |  |
| *Mild (5-9)* | 281 | 36.2 | 1184 | 40.7 | 6 | 40.0 | 1471 | 39.8 |  |  |  |
| *Medium (10-14)* | 116 | 14.9 | 586 | 20.1 | 3 | 20.0 | 705 | 19.1 | 6.1<sup>a</sup> | 10.2<sup>a</sup> | 8.1<sup>a</sup> |
| *Severe (15-19)* | 43 | 5.5 | 276 | 9.5 | 2 | 13.3 | 321 | 8.7 |  |  |  |
| *Most severe (20-27)* | 12 | 1.5 | 119 | 4.1 | 1 | 6.7 | 132 | 3.56 |  |  |  |
| **Panic disorder (PHQ-D)** |  |  |  |  |  |  |  |  |  |  |  |
| *Diagnostic criteria not fulfilled* | 753 | 97.0 | 2722 | 93.6 | 14 | 93.3 | 3489 | 94.3 |  |  |  |
| *Diagnostic criteria fulfilled* | 23 | 3.0 | 187 | 6.4 | 1 | 6.7 | 211 | 5.7 | 1.2<sup>b</sup> | 2.8<sup>b</sup> | 2.0<sup>b</sup> |
| **Other anxiety disorder (PHQ-D)** |  |  |  |  |  |  |  |  |  |  |  |
| *Diagnostic criteria not fulfilled* | 751 | 96.8 | 2662 | 91.5 | 14 | 93.3 | 3427 | 92.6 |  |  |  |
| *Diagnostic criteria fulfilled* | 25 | 3.2 | 247 | 8.5 | 1 | 6.7 | 273 | 7.4 | 1.5<sup>b</sup> | 2.9<sup>b</sup> | 2.2<sup>b</sup> |
| **Insomnia (PSQI)** |  |  |  |  |  |  |  |  |  |  |  |
| *Not fulfilling symptoms (0-5)* | 392 | 50.5 | 1174 | 40.4 | 5 | 33.3 | 1544 | 41.7 |  |  |  |
| *Fulfilling symptoms (6-21)* | 384 | 49.5 | 1735 | 59.6 | 10 | 66.7 | 2156 | 58.3 | 3.8<sup>c</sup> | 7.7<sup>c</sup> | 5.9<sup>c</sup> |

Note. <sup>a</sup> Busch et al. (2013). <sup>b</sup> Jacobi et al. (2014). <sup>c</sup> Schlack, Hapke, Maske, Busch, and Cohrs (2013).

### 3.4 Prediction of mental health symptoms by social support, exercise and exercise specific health competence

Analysis 1 included depressive symptoms as the criterion. Analysis 1 resulted in significant main effects of age, female gender, suffering from a chronic disease, change in employment situation, financial worries, worries about the supply situation, satisfaction with the social life, daily structure, social support, change in exercise, and PA-related affect regulation. The overall model fit was *R*^2^_adjusted_ = 0.41. Results of the analysis are reported in Table 3.

**Table 3:** Results of analysis 1, with depressive symptoms as the criterion variable

| Predictor | *B* | SE | *beta* | *t* | *p*-value | *F* | *df* | *p* | $R^2_{\text{adjusted}}$ |
| --- | --- | --- | --- | --- | --- | --- | --- | --- | --- |
| (Intercept) | 19.33 | 0.59 |  | 32.91 | 0.000 |  |  |  |  |
| Age | -0.06 | 0.01 | -0.14 | -10.39 | 0.000 |  |  |  |  |
| Gender = female<sup>1</sup> | 1.40 | 0.17 | 0.11 | 8.40 | 0.000 |  |  |  |  |
| Gender = diverse<sup>1</sup> | 0.16 | 1.05 | 0.00 | 0.16 | 0.877 |  |  |  |  |
| Relationship Status (single) | 0.25 | 0.14 | 0.02 | 1.79 | 0.073 |  |  |  |  |
| Suffering from Chronic Disease | 2.33 | 0.24 | 0.13 | 9.68 | 0.000 |  |  |  |  |
| Change in Employment Situation | 0.01 | 0.00 | 0.04 | 3.20 | 0.001 |  |  |  |  |
| Financial Worries | 0.02 | 0.00 | 0.13 | 9.47 | 0.000 |  |  |  |  |
| Worries about Supply Situation | 0.02 | 0.00 | 0.06 | 4.62 | 0.000 |  |  |  |  |
| Satisfaction with Social Life | -0.03 | 0.00 | -0.18 | -13.19 | 0.000 |  |  |  |  |
| Daily Structure | -0.04 | 0.00 | -0.23 | -16.36 | 0.000 |  |  |  |  |
| Social Support | -1.64 | 0.10 | -0.22 | -15.71 | 0.000 |  |  |  |  |
| Change in Exercise | -0.25 | 0.04 | -0.08 | -5.69 | 0.000 |  |  |  |  |
| Exercise (min/week)<sup>°</sup> | -0.02 | 0.08 | 0.00 | -0.18 | 0.857 |  |  |  |  |
| PA-related affect regulation<sup>#</sup> | -0.53 | 0.09 | -0.08 | -5.70 | 0.000 |  |  |  |  |
| Exercise (minutes/week) x PA-related affect regulation | -0.13 | 0.10 | -0.02 | -1.29 | 0.198 |  |  |  |  |
|  |  |  |  |  |  | 171.58 | 15.00 | 0.000 | 0.41 |

*Note.* *B*: unstandardized regression weights, *beta*: standardized regression weights; *df*: degrees of freedom; SE: standard error
<sup>1</sup>Reference Category: Gender = male
<sup>°</sup>z-standardized
<sup>#</sup>centered

Depressive symptoms differed statistically significant between the different levels (negative change, no change, positive change) of change in exercise (*F*(2, 3697) = 42.50, *p* < .001). Post-Hoc comparisons indicated, that the mean score for participants, that reported a negative change (*M* = 9.00; *SD* = 5.56) was significantly different to participants that reported no change (*M* = 7.26; *SD* = 5.14) and to participants who reported positive change (*M* = 7.92; *SD* = 5.21). There was no significant difference between participants that reported no change and positive change.

Analysis 2 included anxiety symptoms as the criterion. Analysis 2 resulted in significant main effects of female gender, relationship status, suffering from a chronic disease, change in employment situation, financial worries, worries about the supply situation, satisfaction with the social, daily structure, social support, change in exercise, and PA-related affect regulation. The overall model fit was *R*^2^_adjusted_ = 0.20. Results of the analysis are reported in Table 4.

**Table 4:** Results of analysis 2, with anxiety symptoms as the criterion variable

| Predictor | *B* | SE | *beta* | *t* | *p*-value | *F* | *df* | *p* | $R^2_{\text{adjusted}}$ |
| --- | --- | --- | --- | --- | --- | --- | --- | --- | --- |
| (Intercept) | 1.24 | 0.09 |  | 14.47 | 0.000 |  |  |  |  |
| Age | 0.00 | 0.00 | -0.02 | -1.29 | 0.196 |  |  |  |  |
| Gender = female<sup>1</sup> | 0.23 | 0.02 | 0.14 | 9.27 | 0.000 |  |  |  |  |
| Gender = diverse<sup>1</sup> | 0.09 | 0.15 | 0.01 | 0.61 | 0.543 |  |  |  |  |
| Relationship Status (single) | -0.05 | 0.02 | -0.04 | -2.50 | 0.013 |  |  |  |  |
| Suffering from Chronic Disease | 0.21 | 0.04 | 0.09 | 6.05 | 0.000 |  |  |  |  |
| Change in Employment Situation | 0.00 | 0.00 | 0.04 | 2.36 | 0.018 |  |  |  |  |
| Financial Worries | 0.00 | 0.00 | 0.17 | 10.19 | 0.000 |  |  |  |  |
| Worries about Supply Situation | 0.00 | 0.00 | 0.10 | 6.49 | 0.000 |  |  |  |  |
| Satisfaction with Social Life | 0.00 | 0.00 | -0.14 | -9.09 | 0.000 |  |  |  |  |
| Daily Structure | 0.00 | 0.00 | -0.11 | -6.67 | 0.000 |  |  |  |  |
| Social Support | -0.10 | 0.02 | -0.10 | -6.46 | 0.000 |  |  |  |  |
| Change in Exercise | -0.02 | 0.01 | -0.05 | -2.85 | 0.004 |  |  |  |  |
| Exercise (min/week)<sup>°</sup> | 0.01 | 0.01 | 0.01 | 0.52 | 0.606 |  |  |  |  |
| PA-related affect regulation<sup>#</sup> | -0.03 | 0.01 | -0.04 | -2.40 | 0.016 |  |  |  |  |
| Exercise (minutes/week) x PA-related affect regulation | -0.03 | 0.02 | -0.03 | -1.85 | 0.064 |  |  |  |  |
|  |  |  |  |  |  | 62.79 | 15.00 | 0.000 | 0.20 |

*Note.* *B*: unstandardized regression weights, *beta*: standardized regression weights; *df*: degrees of freedom; SE: standard error
<sup>1</sup>Reference Category: Gender = male
<sup>°</sup>z-standardized
<sup>#</sup>centered

There was a statistically significant difference between participants who reported negative change, no change and positive change in anxiety symptoms (*F*(2, 3697) = 15.52, *p* < .001). Post-Hoc comparisons indicated, that the mean score for participants, that reported a negative change (*M* = 0.86; *SD* = 0.68) was significantly different to participants that reported no change (*M* = 0.73; *SD* = 0.65) and to participants who reported positive change (*M* = 0.73; *SD* = 0.63). There was no significant difference between participants that reported no change and positive change.

Analysis 3 included sleeping disorder symptoms as the criterion. Analysis 3 resulted in significant main effects of female gender, relationship status, suffering from a chronic disease, financial worries, worries about the supply situation, satisfaction with the social life, daily structure, social support, change in exercise, and PA-related affect regulation. The overall model fit was *R*^2^_adjusted_ = 0.25. Results of the analysis are reported in Table 5.

**Table 5:** Results of analysis 3, with sleeping disorder symptoms as the criterion variable

| Predictor | *B* | SE | *beta* | *t* | *p*-value | *F* | *df* | *p* | $R^2_{\text{adjusted}}$ |
| --- | --- | --- | --- | --- | --- | --- | --- | --- | --- |
| (Intercept) | 11.82 | 0.52 |  | 22.63 | 0.000 |  |  |  |  |
| Age | 0.00 | 0.01 | 0.01 | 0.62 | 0.534 |  |  |  |  |
| Gender = female<sup>1</sup> | 0.97 | 0.15 | 0.10 | 6.53 | 0.000 |  |  |  |  |
| Gender = diverse<sup>1</sup> | 0.92 | 0.93 | 0.01 | 0.98 | 0.326 |  |  |  |  |
| Relationship Status (single) | 0.55 | 0.13 | 0.06 | 4.31 | 0.000 |  |  |  |  |
| Suffering from Chronic Disease | 2.24 | 0.21 | 0.15 | 10.46 | 0.000 |  |  |  |  |
| Change in Employment Situation | 0.00 | 0.00 | 0.01 | 0.94 | 0.345 |  |  |  |  |
| Financial Worries | 0.02 | 0.00 | 0.12 | 7.89 | 0.000 |  |  |  |  |
| Worries about Supply Situation | 0.01 | 0.00 | 0.05 | 3.48 | 0.001 |  |  |  |  |
| Satisfaction with Social Life | -0.02 | 0.00 | -0.13 | -8.53 | 0.000 |  |  |  |  |
| Daily Structure | -0.02 | 0.00 | -0.17 | -10.42 | 0.000 |  |  |  |  |
| Social Support | -0.89 | 0.09 | -0.15 | -9.55 | 0.000 |  |  |  |  |
| Change in Exercise | -0.18 | 0.04 | -0.07 | -4.54 | 0.000 |  |  |  |  |
| Exercise (min/week)<sup>°</sup> | -0.14 | 0.07 | -0.03 | -1.86 | 0.063 |  |  |  |  |
| PA-related affect regulation<sup>#</sup> | -0.35 | 0.08 | -0.07 | -4.21 | 0.000 |  |  |  |  |
| Exercise (minutes/week) x PA-related affect regulation | 0.00 | 0.09 | 0.00 | 0.02 | 0.987 |  |  |  |  |
|  |  |  |  |  |  | 84.14 | 15.00 | 0.000 | 0.25 |

*Note.* *B*: unstandardized regression weights, *beta*: standardized regression weights; *df*: degrees of freedom; SE: standard error
<sup>1</sup>Reference Category: Gender = male
<sup>°</sup>z-standardized
<sup>#</sup>centered

Sleeping disorder symptoms differed statistically significant between the different levels (negative change, no change, positive change of change) of change in exercise (*F*(2, 3697) = 39.174, *p* <0.001). Post-Hoc comparisons indicated, that the mean score for participants, that reported a negative change (*M* = 7.93; *SD* = 4.35) was significantly different to participants that reported no change (*M* =6.88; *SD* = 4.16), and to participants who reported positive change (*M* = 6.58; *SD* = 3.76). There was no significant difference between participants that reported no change and positive change.

## 4. Discussion

In this cross-sectional study we examined if perceived social support, exercise in minutes per week and change in exercise are protective factors regarding symptoms of depression, anxiety and sleeping disorder. We further hypothesized that specific exercise related regulation competences enhance the mental health benefits of exercise.

Our descriptive comparison of the prevalence of depression, panic disorder, other anxiety disorder and sleeping disorders resulted in higher prevalence rates during the Covid-19 pandemic in comparison to representative studies conducted before the pandemic. The prevalence needs to be regarded with caution since diagnoses were derived from questionnaires, not delivered by experienced clinicians.

There was no significant association between exercise in minutes and depressive, anxiety, and sleeping disorder symptoms. However, the change in exercise was significantly associated with symptom severity in all three analyses. Exploratory post-hoc analyses resulted in higher symptom severity for participants who decreased their exercise activity in comparison to participants with no change or positive change in exercise activity levels for depression, anxiety and sleeping disorder symptoms.

In line with our hypotheses, perceived social support and PA-related affect regulation was negatively associated with symptoms of anxiety, depression and sleeping disorders. However, we did not find a significant interaction effect between exercise and PA-related affect regulation in any analyses.

In addition, our analyses resulted in significant positive associations between female gender, suffering from a chronic disease, financial worries, and worries about the supply situation with depressive, anxiety, and sleeping disorder symptoms. Satisfaction with the social life and daily structure were negatively associated with depressive, anxiety and sleeping disorder symptoms. Age was negatively associated with depressive symptoms. Change in employment situation was positively associated with depressive and anxiety symptoms. Relationship status was negatively associated with anxiety, but positively associated with sleeping disorder symptoms.

The current cross-sectional survey complements recent studies that reported associations between a decrease in the amount of exercise after the Covid-19 pandemic and higher depression rates (Meyer et al., 2020; Stanton et al., 2020). While Schuch and colleagues (2020) reported, that the effective amount of moderate-to-vigorous exercise was associated with less depression and anxiety disorders, our results indicate only changes in exercise to be associated with mental health benefits. The compelling effects of perceived social support and daily structure on mental health in our analysis complements research, that shows increases in depression and anxiety with the loss of day structure.

Our study has several limitations: Since the design was cross-sectional, results cannot be interpreted causally. Further longitudinal data that will be available in the upcoming measurement will shed more light on the associations between changes in exercise and social support on the attenuation or aggravation of psychopathological symptoms. The sample was mostly composed of females. Further the mean age was quite low (mean age 33 year). Therefore, the representativeness of this sample is limited. Although the recruiting was not directed to people with mental disorders (such as clinics or hospitals) and it was announced as a study about the “protective factors for mental health”, there might be a self-selection bias. Especially people that suffered from mental problem might have been more interested in participating in the study causing the very high prevalence. Further methodological limitations comprise the assessment of the change in exercise and anxiety symptoms (only one item each). Further work-related and leisure-time related PA was not analyzed, which could bias our results. Lastly the model fit can be assumed as adequate for depression with an explained variance of 41 %, however the explained variance for anxiety (20 %) and sleep (26 %) can be considered as low.

## Data Availability

The data that support the findings of this study are available upon reasonable request from the corresponding author. The data are not publicly available due to the third point of measurement being not yet completed.

## Acknowledgements

This research has not been funded by an external source. We thank all the participants who took time to take part in this research project. Particularly, we want to thank the podcast “Lage der Nation” who mentioned the study in their podcast episode.

## Authors contributions

Study conception and design: Leonie Louisa Bauer, Britta Seiffer, Sebastian Wolf, Clara Deinhart, Beatrice Atrott, Gorden Sudeck, Martin Hautzinger. Acquisition of data: Leonie Louisa Bauer, Britta Seiffer, Sebastian Wolf, Clara Deinhart, Beatrice Atrott. Analysis and interpretation of data: Sebastian Wolf, Britta Seiffer, Clara Deinhart, Beatrice Atrott, Leonie Louisa Bauer, Inka Rösel. Drafting of manuscript: Leonie Louisa Bauer, Britta Seiffer, Sebastian Wolf. Critical revision: All authors.

## Notes

### Competing Interest Statement

The authors have declared no competing interest.

### Clinical Trial

The study was registered in German Clinical Trial Register (DRKS00021791). To be able to assess the effects of the Covid-19 measures (lock down, social distancing) we needed start the assessments prior to the study registration, which was 4 weeks after the first measurement point. Data needed to be collected as soon as possible. However the information provided in the trial registration does not differ from the information in the application for the ethical approval, which was obtained prior to the inclusion of the first participant. The application of the ethical approval can be accessed on request.

### Funding Statement

This study was not funded by external sources.

### Author Declarations

The study was approved by the local ethics committee for research at the Faculty of Economics and Social Sciences.

## References

American College of Sports Medicine. (2017). ACSM’s Guidelines for Exercise Testing and Prescription. Alphen aan den Rijn: Wolters Kluwer.

Brooks, S. K., Webster, R. K., Smith, L. E., Woodland, L., Wessely, S., Greenberg, N., & Rubin, G. J. (2020). The psychological impact of quarantine and how to reduce it: rapid review of the evidence. The Lancet, 395(10227), 912–920. doi:10.1016/S0140-6736(20)30460-8

Busch, M. A., Maske, U. E., Ryl, L., Schlack, R., & Hapke, U. (2013). Prävalenz von depressiver Symptomatik und diagnostizierter Depression bei Erwachsenen in Deutschland [Prevalence of depressive symptoms and diagnosed depression in adults in Germany]. Bundesgesundheitsblatt-Gesundheitsforschung-Gesundheitsschutz, 56(5-6), 733–739. doi:10.1007/s00103-013-1688-3

Buysse, D. J., Reynolds, C. F., 3rd, Monk, T. H., Berman, S. R., & Kupfer, D. J. (1989). The Pittsburgh Sleep Quality Index: a new instrument for psychiatric practice and research. Psychiatry Res, 28(2), 193–213. doi:10.1016/0165-1781(89)90047-4

Cao, W., Fang, Z., Hou, G., Han, M., Xu, X., Dong, J., & Zheng, J. (2020). The psychological impact of the COVID-19 epidemic on college students in China. Psychiatry Res, 287, 112934. doi:10.1016/j.psychres.2020.112934

Carl, J., Sudeck, G., & Pfeifer, K. (2020). Competencies for a Healthy Physically Active Lifestyle— Reflections on the Model of Physical Activity-Related Health Competence. J Phys Act Health, 1(aop), 1–10.

Dalgard, O. S., Dowrick, C., Lehtinen, V., Vazquez-Barquero, J. L., Casey, P., Wilkinson, G., … Group, O. (2006). Negative life events, social support and gender difference in depression: A multinational community survey with data from the ODIN study. Soc Psychiatry Psychiatr Epidemiol, 41(6), 444–451. doi:10.1007/s00127-006-0051-5

Doi, Y., Minowa, M., Uchiyama, M., & Okawa, M. (2001). Subjective sleep quality and sleep problems in the general Japanese adult population. Psychiat Clin Neurosci, 55(3), 213–215. doi:10.1046/j.1440-1819.2001.00830.x

Federal Employment Agency. (2020). Auswirkungen der Corona-Krise auf den Arbeitsmarkt [Effects of the Corona crisis on the labour market]. Berichte: Arbeitsmarkt kompakt - Auswirkungen der Corona-Krise. Retrieved from https://statistik.arbeitsagentur.de/Statistikdaten/Detail/202005/arbeitsmarktberichte/am-kompakt-corona/am-kompakt-corona-d-0-202005-pdf.pdf

Fuchs, R., Klaperski, S., Gerber, M., & Seelig, H. (2015). Messung der Bewegungs-und Sportaktivität mit dem BSA-Fragebogen. Z Gesundheitspsychol, 23(2), 60–76. doi:10.1026/0943-8149/a000137

Fydrich, T., Sommer, G., Tydecks, S., & Brähler, E. (2009). Fragebogen zur sozialen Unterstützung (F-SozU): Normierung der Kurzform (K-14). Z Med Psychol, 18, 43–48.

Gariepy, G., Honkaniemi, H., & Quesnel-Vallee, A. (2016). Social support and protection from depression: systematic review of current findings in Western countries. Brit J Psychiatry, 209(4), 284–293. doi:10.1192/bjp.bp.115.169094

Gräfe, K., Zipfel, S., Herzog, W., & Löwe, B. (2004). Screening psychischer Störungen mit dem “Gesundheitsfragebogen für Patienten (PHQ-D)”. Diagnostica, 50(4), 171–181. doi:10.1026/0012-1924.50.4.171

Grav, S., Hellzen, O., Romild, U., & Stordal, E. (2012). Association between social support and depression in the general population: the HUNT study, a cross-sectional survey. J Clin Nurs, 21(1-2), 111–120. doi:10.1111/j.1365-2702.2011.03868.x

Harvey, S. B., Overland, S., Hatch, S. L., Wessely, S., Mykletun, A., & Hotopf, M. (2018). Exercise and the Prevention of Depression: Results of the HUNT Cohort Study. Am J Psychiatry, 175(1), 28–36. doi:10.1176/appi.ajp.2017.16111223

Huang, Y., & Zhao, N. (2020). Generalized anxiety disorder, depressive symptoms and sleep quality during COVID-19 outbreak in China: a web-based cross-sectional survey. Psychiatry Res, 288, 112954. doi:10.1016/j.psychres.2020.112954

Jacobi, F., Hofler, M., Strehle, J., Mack, S., Gerschler, A., Scholl, L., … Wittchen, H. U. (2014). [Mental disorders in the general population : Study on the health of adults in Germany and the additional module mental health (DEGS1-MH)]. Nervenarzt, 85(1), 77–87. doi:10.1007/s00115-013-3961-y

Lee, A. M., Wong, J. G., McAlonan, G. M., Cheung, V., Cheung, C., Sham, P. C., … Chua, S. E. (2007). Stress and psychological distress among SARS survivors 1 year after the outbreak. Can J Psychiatry, 52(4), 233–240. doi:10.1177/070674370705200405

Leiner, D. (2016). SoSci Survey [Computer Software] (Version 2.6.00). Retrieved from https://www.soscisurvey.de

Liu, C. H., Zhang, E., Wong, G. T. F., & Hyun, S. (2020). Factors associated with depression, anxiety, and PTSD symptomatology during the COVID-19 pandemic: Clinical implications for US young adult mental health. Psychiatry Res, 113172. doi:10.1016/j.psychres.2020.113172

Löwe, B., Spitzer, R. L., Zipfel, S., & Herzog, W. (2002). PHQ-D. Gesundheitsfragebogen für Patienten. Manual. Komplettversion und Kurzform. Autorisierte deutsche Version des „Prime MD Patient Health Questionnaire (PHQ)”. Karlsruhe: Pfizer.

Meyer, J., McDowell, C., Lansing, J., Brower, C., Smith, L., Tully, M., & Herring, M. (2020). Changes in physical activity and sedentary behaviour due to the COVID-19 outbreak and associations with mental health in 3,052 US adults. Cambridge Open Engage. doi:10.33774/coe-2020-h0b8g

Miloyan, B., Joseph Bienvenu, O., Brilot, B., & Eaton, W. W. (2018). Adverse life events and the onset of anxiety disorders. Psychiatry Res, 259, 488–492. doi:10.1016/j.psychres.2017.11.027

Paul, K. I., Geithner, E., & Moser, K. (2009). Latent deprivation among people who are employed, unemployed, or out of the labor force. J Psychol, 143(5), 477–491. doi:10.3200/JRL.143.5.477-491

Rossi, R., Socci, V., Talevi, D., Mensi, S., Niolu, C., Pacitti, F., … Di Lorenzo, G. (2020). COVID-19 pandemic and lockdown measures impact on mental health among the general population in Italy. An N=18147 web-based survey. medRxiv, 2020.2004.2009.20057802. doi:10.1101/2020.04.09.20057802

Santini, Z. I., Jose, P. E., Cornwell, E. Y., Koyanagi, A., Nielsen, L., Hinrichsen, C., … Koushede, V. (2020). Social disconnectedness, perceived isolation, and symptoms of depression and anxiety among older Americans (NSHAP): a longitudinal mediation analysis. Lancet Public Health, 5(1), E62–E70. doi:10.1016/S2468-2667(19)30230-0

Schlack, R., Hapke, U., Maske, U., Busch, M., & Cohrs, S. (2013). Frequency and distribution of sleep problems and insomnia in the adult population in Germany: results of the German Health Interview and Examination Survey for Adults (DEGS1). Bundesgesundheitsblatt Gesundheitsforschung Gesundheitsschutz, 56(5-6), 740–748. doi:10.1007/s00103-013-1689-2

Schuch, F. B., Bulzing, R., Meyer, J., Vancampfort, D., Firth, J., Stubbs, B., … Smith, L. (2020). Associations of moderate to vigorous physical activity and sedentary behavior with depressive and anxiety symptoms in self-isolating people during the COVID-19 pandemic: A cross-sectional survey in Brazil. Scielo Preprints. doi:10.1590/SciELOPreprints.526

Schuch, F. B., Stubbs, B., Meyer, J., Heissel, A., Zech, P., Vancampfort, D., … Hiles, S. A. (2019). Physical activity protects from incident anxiety: A meta-analysis of prospective cohort studies. Depress Anxiety, 36(9), 846–858. doi:10.1002/da.22915

Schuch, F. B., Vancampfort, D., Firth, J., Rosenbaum, S., Ward, P. B., Silva, E. S., … Stubbs, B. (2018). Physical Activity and Incident Depression: A Meta-Analysis of Prospective Cohort Studies. Am J Psychiatry, 175(7), 631–648. doi:10.1176/appi.ajp.2018.17111194

Stanton, R., To, Q. G., Khalesi, S., Williams, S. L., Alley, S. J., Thwaite, T. L., … Vandelanotte, C. (2020). Depression, Anxiety and Stress during COVID-19: Associations with Changes in Physical Activity, Sleep, Tobacco and Alcohol Use in Australian Adults. Int J Environ Res Public Health, 17(11). doi:10.3390/ijerph17114065

Sudeck, G., Jeckel, S., & Schubert, T. (2018). Individual Differences in the Competence for Physical-Activity-Related Affect Regulation Moderate the Activity-Affect Association in Real-Life Situations. J Sport Exerc Psychol, 40(4), 196–205. doi:10.1123/jsep.2018-0017

Sudeck, G., & Pfeifer, K. (2016). Physical activity-related health competence as an integrative objective in exercise therapy and health sports - conception and validation of a short questionnaire. Ger J Exerc Sport Res, 46(2), 74–87. doi:10.1007/s12662-016-0405-4

Wang, C., Pan, R., Wan, X., Tan, Y., Xu, L., Ho, C. S., & Ho, R. C. (2020). Immediate Psychological Responses and Associated Factors during the Initial Stage of the 2019 Coronavirus Disease (COVID-19) Epidemic among the General Population in China. Int J Environ Res Public Health, 17(5). doi:10.3390/ijerph17051729

Wang, X., Cai, L., Qian, J., & Peng, J. (2014). Social support moderates stress effects on depression. Int J Ment Health Syst, 8(1), 41. doi:10.1186/1752-4458-8-41

Wittchen, H., Krause, P., Höfler, M., Winter, S., Spiegel, B., Hajak, G., … Pfister, H. (2001). NISAS-2000 – die „Nationwide Insomnia Screening and Awareness Study”. Nervenheilkunde 2001. Nervenheilkunde, 20, 4–16.

World Health Organization. (2020). WHO Coronavirus Disease (COVID-19) Dashboard. Retrieved from https://covid19.who.int/

Xiao, H., Zhang, Y., Kong, D., Li, S., & Yang, N. (2020). The Effects of Social Support on Sleep Quality of Medical Staff Treating Patients with Coronavirus Disease 2019 (COVID-19) in January and February 2020 in China. Med Sci Monit, 26, e923549. doi:10.12659/MSM.923549

Yip, P. S., Cheung, Y. T., Chau, P. H., & Law, Y. W. (2010). The impact of epidemic outbreak: the case of severe acute respiratory syndrome (SARS) and suicide among older adults in Hong Kong. Crisis, 31(2), 86–92. doi:10.1027/0227-5910/a000015

